# Efficacy of the AZD1222 (ChAdOx1 nCoV-19) COVID-19 Vaccine Against SARS-CoV-2 Variants of Concern

**DOI:** 10.1101/2022.07.18.22277743

**Authors:** Anthonet L. Koen, Alane Izu, Vicky Baillie, Gaurav Kwatra, Clare L. Cutland, Lee Fairlie, Sherman D. Padayachee, Keertan Dheda, Shaun L. Barnabas, Qasim Ebrahim Bhorat, Carmen Briner, Khatija Ahmed, Sutika Bhikha, Jinal N. Bhiman, Jeanine du Plessis, Aliasgar Esmail, Elizea Horne, Shi-Hsia Hwa, Aylin Oommen-Jose, Teresa Lambe, Matt Laubscher, Mookho Malahleha, Gabriella Benade, Shakeel McKenzie, Suzette Oelofse, Faeezah Patel, Sureshnee Pillay, Sarah Rhead, Hylton Rodel, Carol Taoushanis, Houriiyah Tegally, Asha Thombrayil, Tonya L. Villafana, Sarah Gilbert, Andrew J. Pollard, Shabir A. Madhi

## Abstract

In this South African phase 1/2b study, we demonstrated vaccine efficacy (VE) of two doses of AZD1222 for asymptomatic and symptomatic SARS-CoV-2 infection: 90.6% against wild-type and 77.1% against the Delta variant ≥9 months after vaccination. VE against infection with the Beta variant, which preceded circulation of Delta, was 6.7%.

**Clinical trial identifier:** CT.gov NCT04444674

## Introduction

Coronavirus Disease 2019 (COVID-19) vaccines have been rapidly developed and approved for licensure over the past two years. The emergence of new variants of concern (VoCs) containing mutations that affect transmissibility or confer the ability to evade vaccine- and infection-induced antibodies requires analysis of vaccine efficacy (VE) against specific SARS-CoV-2 variants.

South Africa experienced an ancestral SARS-CoV-2 (wild type, WT) wave in March 2020, which peaked in July 2020 [1]. The Beta (B.1.351) variant, which possessed relative neutralization resistance to antibodies induced by ancestral virus infection and most first-generation COVID-19 vaccines, fueled a second wave of infections from November 2020, peaking in January 2021. This was followed by a third wave from July–September 2021 driven by infections caused by the Delta variant [2], which was two-fold more transmissible than the ancestral virus.

An interim analysis using data from four AZD1222 (ChAdOx1 nCoV-19) studies conducted in Brazil, South Africa and the UK between April 23, 2020–November 4, 2020, reported an overall VE for symptomatic infection of 66.7% (95% confidence interval [CI], 57.4%□74.0%) [3]. VE at preventing symptomatic infection with the Alpha (B.1.1.7) variant in the UK was 74.6% (95% CI, 41.6%□88.9%) [4]. Primary analysis of the pivotal phase 3 trial of AZD1222 demonstrated a VE of 74.0% (95% CI, 65.3%□80.5%) at preventing symptomatic COVID-19 with predominantly the ancestral virus in the United States, Chile, and Peru (data cut-off: March 5, 2021) [5].

Previously, we reported an overall VE for two-dose primary series AZD1222 vaccination of 21.9% (95% CI, -49.9%□59.8%) against mild-to-moderate COVID-19 in SARS-CoV-2-naïve South African participants (data-cut off: January 15, 2021) [6]. The majority of these cases were driven by the Beta variant with a VE of 10.4% (95% CI, -76.8%□54.8%) against mild-to-moderate COVID-19 observed in a secondary analysis [6]. In this brief report, we report our final analysis for overall and variant-specific VE and safety of primary series AZD1222 from a multicenter phase 1b/2 trial in South Africa.

## Methods

### Trial Design

Full details of this phase 1b/2 double-blind, randomized, placebo-controlled study (NCT04444674; PACTR202006922165132) assessing the safety and efficacy of AZD1222 in South African adults have been reported previously [6]. Healthy adults aged 18–65 years were recruited between June 24, 2020–November 9, 2020 and randomized to receive two doses of placebo (saline) or AZD1222 in a 1:1 ratio. This is the final VE analysis through to the time of unblinding of participants.

### Endpoints

The primary endpoint was VE in the context of polymerase chain reaction (PCR)-positive symptomatic COVID-19 occurring >14 days after the second dose of study drug, and safety. Secondary objectives included VE based on SARS-CoV-2 variant, and reactogenicity.

### Laboratory Methods

Laboratory methods are detailed in **Supplementary Methods**.

### Statistical Methods

Statistical methods for COV005 have been published previously [6]. Descriptions of the statistical methods used in this report are detailed in **Supplementary Methods**.

### Ethics Statement

This study was conducted in accordance with the principles outlined in the Declaration of Helsinki and Good Clinical Practice guidelines. Participants were required to provide informed consent. The study was approved by the South African Health Products Regulatory Authority and institutional Ethics Committees.

## Results

2130 healthy adults (including 104 living with HIV) aged 18–65 were randomized to receive two doses of placebo (1065; 50%) or AZD1222 (1065; 50%). A total of 960 (placebo) and 935 (vaccinated) participants were included in this analysis (Supplementary Figure 1). 107 (31 randomized to placebo and 76 randomized to AZD1222) participants did not receive two doses of the allocated treatment.126 participants were infected by SARS-CoV-2 <14 days post-second dose (72 placebo and 54 AZD1222) and two placebo participants died before <14 days post-second dose. The median age of participants was 31 years, with most participants aged 18–45 (82.7%; Supplementary Table 1); 49.8% of participants were of normal weight and 42.6% were smokers. 7.5% of participants had either hypertension, diabetes or medical conditions related to the respiratory system (Supplementary Table 1).

The increased frequency of Beta, Delta and other VoCs temporally coincided with the peaks of infection in the study cohort (Supplementary Figure 2). The first wave of South African infections (June–August 2020) was dominated by the circulating WT, whereas the second (November 2020–March 2021) and third (May–August 2021) waves were mainly dominated by Beta and Delta variants, respectively.

### Vaccine Efficacy

For any infection, the mean time between second study injection and SARS-CoV-2 infection was 5.5 months in the placebo group and 4.5 months in vaccinees (Supplementary Table 2); for the Delta variant, the mean time was 9.3 and 10.0 months, respectively. Follow-up time post second study injection was similar for mild and moderate cases.

When examining the effect on SARS-CoV-2 infection (i.e., inclusive of asymptomatic infection and cases not meeting criteria of severity), overall VE against SARS-CoV-2 infection was 38.3% (95% CI, 15.1%□55.4%; Supplementary Table 2). VE against infection with WT was 90.6% (95% CI, 35.4%□99.8%) and 77.1% (95% CI, 30.4%□94.4%) against the Delta variant; whilst there was lack of VE against the Beta variant (6.7%; 95% CI, - 41.1%□38.5%). Cumulative incidence over time for SARS-CoV-2 infection due to WT, Beta and Delta variants is shown in Figure 1.

**Figure 1.**
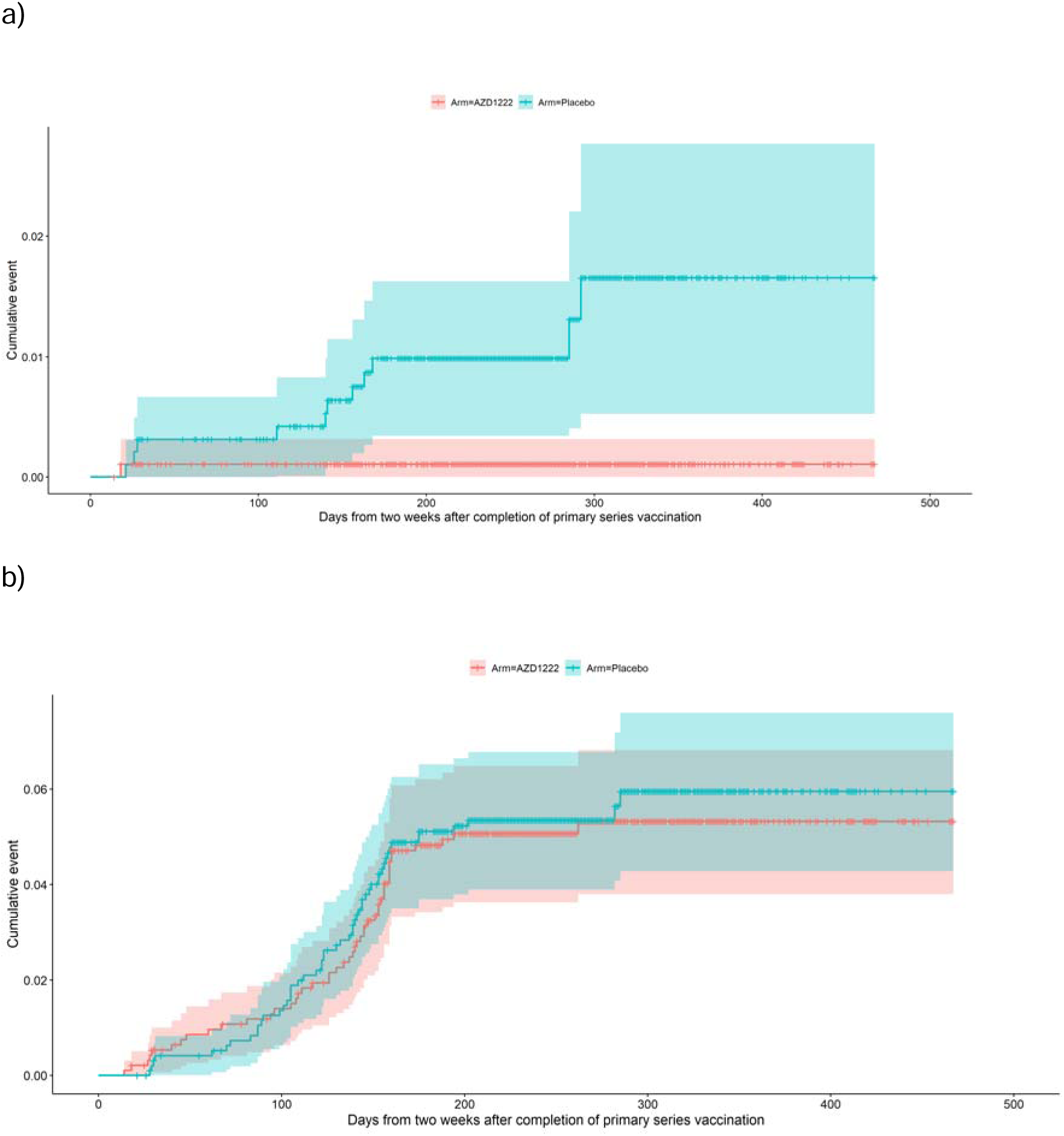
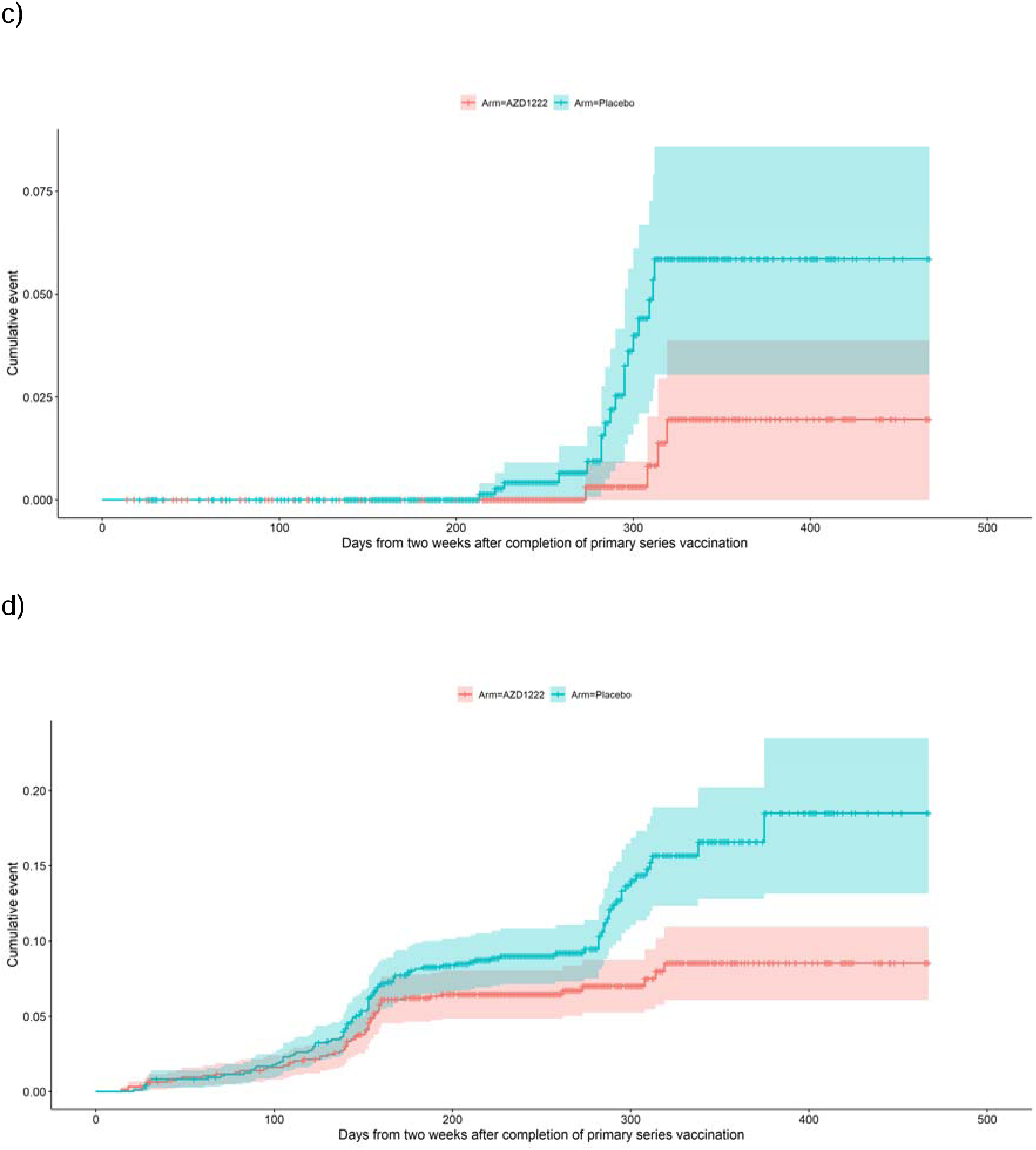
Cumulative incidence over time for any SARS-CoV-2 infection (including both asymptomatic and symptomatic illness) due to wild type (a), Beta (b) and Delta (c) variants, and overall (d).

Overall VE against mild and moderate COVID-19 (no severe cases documented in the cohort before unblinding) for any variant was 45.2% (95% CI, 19.7%□63.1%; Supplementary Table 2). VE varied by variant: 88.5% (95% CI, 17.2%□99.7%) against WT and 75.7% (95% CI, 25.6%□94.1%) against the Delta variant; VE against the Beta variant was 8% (95% CI, - 51.3%□44.2%). Similar VE point-estimates were evident for only moderate COVID-19 although the CIs were wider. Distribution of infections by vaccination status, variant and severity are shown in Supplementary Table 3, and Supplementary Table 4 contextualizes the timing of sequences that were unclassified as part of this analysis. A sensitivity analysis showing the cumulative incidence for infection across variants, accounting for unclassified sequences, is shown in Supplementary Figure 3.

### Adverse Events

Adverse events and severe adverse events are presented in Supplementary Tables 6 and 7, respectively. As previously reported [6], one vaccinee had severe Grade 3 fever which was considered related to AZD1222 by the investigator and was resolved within 48 hours.

## Discussion

Here, we present our final analysis of overall and variant-specific VE and safety of an AZD1222 primary series. We demonstrate high VE against WT SARS-CoV-2 infection, in line with previous findings [3, 5]. VE against the Delta variant was also high, despite infections occurring 9–10 months after study participants had received their second dose of AZD1222. Given the timing of emergence of Delta, the lower VE we observed compared to WT may be due to waning of antibodies, viral escape from neutralizing antibodies induced by vaccination and increased transmissibility of this variant [7]. Despite this, VE observed against Delta across this timeframe points to good durability of protection, likely due to an anamnestic response from AZD1222. It is notable that no cases of severe disease were documented ahead of unblinding.

In contrast to WT and Delta variants, and in line with our previous interim analysis, VE against mild-moderate COVID-19 due to Beta variant was low, which was likely due to escape from vaccine-induced neutralizing antibody activity [6]. However, a subsequent real-world evidence study demonstrated an 83% risk reduction for hospitalization or death due to infection with the Beta or Gamma VoCs following AZD1222 vaccination, suggesting that cellular immunity may continue to provide protection against severe disease despite neutralizing antibody escape [8]. Despite the limited protection against symptomatic disease with Beta, vaccination campaigns that continued to deploy AZD1222 in line with World Health Organization recommendations enabled the prevention of deaths in subsequent waves driven by Delta. The Delta variant wave in South Africa was the most severe in relation to deaths, contributing to 50% of the overall death toll which transpired from COVID-19 since the start of the pandemic until the end of the fourth wave, which was due to the Omicron variant [9]. Overall, this highlights that national immunization campaigns should not discontinue use of a vaccine based on experience with a single variant.

At the time of writing, the Omicron (B.1.1.529) VoC and sub-lineages thereof are the dominant variants across much of the world, with cases of a Delta Omicron (BA.1 x AY.4) recombinant variant also documented [10]. Third dose booster vaccination has been widely used a mitigation strategy for breakthrough Omicron infection. However, due to preliminary immunogenicity studies favoring AZD1222–BNT162b2 or homologous BNT162b2 boosting strategies, booster doses of AZD1222 have largely been restricted to those unable to receive mRNA vaccines. Real-world data has shown similar vaccine effectiveness against symptomatic disease and hospitalization from Omicron with either an AZD1222 or BNT162b2 booster following AZD1222 primary series, indicating the potential wider utility of AZD1222 as a booster [11]. As our analysis was censored and participants unblinded before the emergence of Omicron, we await further studies to determine the VE of AZD1222 and other SARS-CoV-2 vaccines as primary series and boosters against Omicron and related subvariants.

## Data Availability

Data are available at www.wits-vida.org; requests for data sharing should be directed to Professor Shabir A. Madhi

## Acknowledgments

The authors gratefully acknowledge the local safety physician, Guy Richards, for reviewing all SAEs. The authors would like to acknowledge the Data Safety and Monitoring Committee for the trial: Robert Heyderman (co-chair), Manish Sadarangani (co-chair), Paul Kaye, Steve Black, George Bouliotis, Gregory Hussey, Bernhards Ogutu, Walter Orenstein, Sonia Ramos, Cornelia L. Dekker, Elizabeth Bukusi.

The authors are grateful to the Independent Case Evaluation Committee:

Jeremy Carr, Steve Chambers, Kim Davis, Simon Drysdale, Charles Feldman, Malick Gibani, Elizabeth Hammershaimb, Michael Harrington, Celina Jin, Seilesh Kadambari, Rama Kandasamy, Carla Leisegang, Toby Maher, Jamilah Meghji, Marc Mendelson, Colin Menezes, Claire Munro, Jeremy Nel, David Pace, Rekha Rapaka, Robindra Basu Roy, Daniel Silman, Gemma Sinclair, Merika Tsitsi, Jing Wang.

The authors would like to acknowledge trial team staff members for their valued contributions towards the trial.

Medical editing support was provided by Rose Follis, PhD, Craig O’Hare, PhD, and Jon Moran, PhD of Ashfield MedComms, an Ashfield company, and was funded by AstraZeneca in accordance with Good Publication Practice (GPP3) guidelines (Battisti WP, *et al*. Ann Intern Med. 2015;163:461–464).

