## Supplementary Materials for "Efficacy of the AZD1222 (ChAdOx1 nCoV-19) COVID-19 Vaccine Against SARS-CoV-2 Variants of Concern"

**Supplementary Methods**

*Laboratory Methods*

*SARS-CoV-19 nucleic acid amplification test (NAAT) testing*

All NAAT testing was done at one of two study-site laboratories, Vaccines and Infectious Diseases Analytics Research Unit (VIDA, Johannesburg) or University of Cape Town Lung Institute (Cape Town, South Africa). Nasopharyngeal swabs were tested for the presence of SARS-CoV-2 using Emergency Use Authorization assays developed by the Centre for Diseases Control and Prevention (CDC) which target two regions within the nucleocapsid gene (N1 and N2) and a third assay which detects the human RNase P gene [1, 2]. Results were classified as positive for SARS-CoV-2 when both the N1 and N2 targets were detected at a cycle threshold (Ct) <40. Results were classified as inconclusive if only N1 or N2 was detected with Ct values <40. If a repeat swab test tested inconclusive, the participant was considered to be infected with SARS-CoV-2. In order for negative results to be valid, the human RNase P assay needed to be detected with a cycle threshold <40.

*Whole genome sequencing*

Whole genome sequencing of samples was done either at VIDA or the KwaZulu-Natal Research Innovation and Sequencing Platform (KRISP). Superscript IV with random hexamers (Life Technologies, Carlsbad, CA) were used to generate cDNA from SARS-CoV-2 NAAT confirmed NP samples. The QIAseq DIRECT SARS-CoV-2 Primer Panel (QIAGEN, Germany) or the ARCTIC V3 protocol [3-5] and Illumina^®^ Nextera Flex DNA Library Prep kits were used to amplify, and index paired end libraries of genomic DNA according to manufacturers’ instructions [6]. These libraries were sequenced on a 500 cycle v2 MiSeq Reagent Kit on an Illumina MiSeq instrument (Illumina, San Diego, CA, USA).

*Genome assembly and phylogenetic analysis*

Genome Detective 1.126 (https://www.genomedetective.com) and the Coronavirus Typing Tool were used to generate paired-end fastq reads [7]. Low-quality mutations were filtered out of the initial assembly generated from Genome Detective using bcftools 1.7-2 mpileup method. The whole genomes were compared against the global reference dataset using a custom pipeline based on a local version of NextStrain for variant classification of sequenced isolates. The pipeline contains several python scripts that manage the analysis workflow. It performs alignment of genotypes in Nextalign (https://github.com/neherlab/nextalign), phylogenetic tree inference in IQ-Tree, tree dating [8] and ancestral state construction and annotation(<https://github.com/nextstrain/ncov>).

*Statistical analyses*

Incidence of COVID-19 disease was calculated as the number of participants with COVID-19 divided by the total in person follow-up time in days. Participants were followed from two weeks post-second dose until the first of the following events: withdrawal, death, SARS-CoV-2 infection, unblinding or censored by 15 November 2021. VE was calculated as 1 minus the relative risk (incidence in vaccinated individuals divided by incidence in placebo recipients) and 95% CI were calculated using the Clopper-Pearson exact methods. Participants were grouped by treatment received (placebo or vaccine), regardless of their planned group assignment. Only participants who received two doses of placebo or vaccine and had sufficient follow-up were included in the analysis. VE was assessed against COVID-19 of any severity, and infection for wild-type (WT), Beta and Delta variants. A sensitivity analysis was performed to include infections where the strain was unclassifiable. Infections pre-October 2020 were assigned a WT classification, infections between October 2020 and April 2021 were assigned Beta, and infections between May and November 2021 were assigned Delta. Local and systemic Adverse events (AEs) occurring in the 28 days following each vaccination were recorded. Serious AEs (SAEs) were collected through the whole study period.

**Supplementary Table 1. Demographics for participants included in the efficacy analysis**

| **Variable** | **Overall n = 1895** | **Placebo**  **N = 960** | **Vaccine**  **N = 935** |
| --- | --- | --- | --- |
| Male, n (%) | 1047 (55.3) | 520 (54.2) | 527 (56.4) |
| HIV positive, n (%) | 98 (5.2) | 48 (5) | 50 (5.3) |
| Median Age, years (IQR) | 31 (25–41) | 31 (25–40) | 31 (25–41) |
| 18–<45 | 1568 (82.7) | 798 (83.1) | 770 (82.4) |
| 45–<60 | 288 (15.2) | 139 (14.5) | 149 (15.9) |
| ≥60 | 39 (2.1) | 23 (2.4) | 16 (1.7) |
| BMI, n (%) | | | |
| Underweight | 148 (7.8) | 66 (6.9) | 82 (8.8) |
| Normal | 942 (49.8) | 483 (50.3) | 459 (49.2) |
| Overweight | 433 (22.9) | 209 (21.8) | 224 (24) |
| Obese | 370 (19.5) | 202 (21) | 168 (18) |
| Smoker, n (%) | 808 (42.6) | 394 (41) | 414 (44.3) |
| Alcohol, n (%) | 931 (49.1) | 469 (48.9) | 462 (49.4) |
| Health worker, n (%) | 157 (8.3) | 87 (9.1) | 70 (7.5) |
| Race, n (%) | | | |
| Black | 1330 (70.3) | 674 (70.4) | 656 (70.2) |
| Mixed | 281 (14.9) | 138 (14.4) | 143 (15.3) |
| White | 244 (12.9) | 128 (13.4) | 116 (12.4) |
| Other | 36 (1.9) | 17 (1.8) | 19 (2) |
| Complications, n (%) | 143 (7.5) | 68 (7.1) | 75 (8) |
| Hypertension | 67 (3.5) | 33 (3.4) | 34 (3.6) |
| Respiratory system | 73 (3.9) | 33 (3.4) | 40 (4.3) |
| Diabetes | 8 (0.4) | 4 (0.4) | 4 (0.4) |
| Time between doses, median days (IQR) | 28 (28–32) | 28 (28–32) | 28 (28–32) |
| Follow-up time, median days (IQR)* | 250 (216–298) | 251 (219–301) | 249 (213–291) |
| Total follow-up time, per person-days* | 480697 | 244140 | 236557 |

Abbreviations: BMI, body mass index; IQR, interquartile range.

*Follow-up time was calculated as the time from 2^nd^ injection to first of the following events: withdrawal, death, SARS-CoV-2 infection, unblinding or censored by 15 November 2021.

.

**Supplementary Table 2. Vaccine efficacy**

| **Severity** | **SARS-CoV-2** | **Total cases** | **Placebo n = 960 Total FU time(days) = 244,140 Mean FU in days (SD) = 254 (71)** | | | | **Vaccine n = 935 Total FU time (days) = 236,557 Mean FU in days (SD) = 253 (74)** | | | **VE  (95% CI)** |
| --- | --- | --- | --- | --- | --- | --- | --- | --- | --- | --- |
|  |  |  | **n (%)** | **Mean FU in months**  **(SD)** | **Incidence rate (person-days)** | **n (%)** | | **Mean FU in months**  **(SD)** | **Incidence rate (person-days)** |  |
| **Any infection*** | Any^†^ | 171 | 107 (11.1) | 5.5 (2.7) | 160 | 64 (6.8%) | | 4.5 (2.2) | 98.7 | 38.3% (15.1, 55.4) |
|  | WT | 12 | 11 (1.1) | 4.6 (3.1) | 16.4 | 1 (0.1%) | | 0.6 (NA) | 1.5 | 90.6% (35.4, 99.8) |
|  | Beta | 99 | 52 (5.4) | 4.2 (1.7) | 77.7 | 47 (5%) | | 4.0 (1.7) | 72.5 | 6.7% (-41.1, 38.5) |
|  | Delta | 22 | 18 (1.9) | 9.3 (1.0) | 26.9 | 4 (0.4%) | | 10.0 (0.7) | 6.2 | 77.1% (30.4, 94.4) |
| **Mild/ Moderate** | Any^†^ | 124 | 81 (8.4) | 5.6 (2.9) | 121.1 | 43 (4.6%) | | 4.4 (2.6) | 66.3 | 45.2% (19.7, 63.1) |
|  | WT | 10 | 9 (0.9) | 4.6 (3.4) | 13.5 | 1 (0.1%) | | 0.6 (NA) | 1.5 | 88.5% (17.2, 99.7) |
|  | Beta | 70 | 37 (3.9) | 4.1 (1.8) | 55.3 | 33 (3.5%) | | 3.8 (1.9) | 50.9 | 8% (-51.3, 44.2) |
|  | Delta | 21 | 17 (1.8) | 9.4 (0.9) | 25.4 | 4 (0.4%) | | 10 (0.7) | 6.2 | 75.7% (25.6, 94.1) |

Abbreviations: CI, confidence interval; FU, follow-up; SD, standard deviation; VE, vaccine efficacy; WT, wild type; NA, not applicable.

^*^Includes symptomatic and asymptomatic infections. ^†^Includes infections from strains that were not classified as WT, Beta or Delta.

**Supplementary Table 3. Variant distribution by severity**

| **Severity** | **Total cases** | **WT** | **Beta** | **Delta** | **Failed library preparation^†^** | **Unclassified^‡^** |
| --- | --- | --- | --- | --- | --- | --- |
| **Any infection*** | 171^†^ | 12 | 99 | 22 | 8 | 30 |
| Placebo | 107 | 11 | 52 | 18 | 6 | 20 |
| Vaccine | 64 | 1 | 47 | 4 | 2 | 10 |
| **Mild/Moderate** | 124 | 10 | 70 | 21 | 5 | 18 |
| Placebo | 81 | 9 | 37 | 17 | 5 | 13 |
| Vaccine | 43 | 1 | 33 | 4 | 0 | 5 |
| **Moderate** | 45 | 7 | 27 | 5 | 2 | 4 |
| Placebo | 27 | 6 | 13 | 4 | 2 | 2 |
| Vaccine | 18 | 1 | 14 | 1 | 0 | 2 |

Abbreviation: WT, wild type.
*Includes symptomatic and asymptomatic infections.

^†^One severe case happened after unblinding.

^‡^See Supplementary Table S4 for further details.

Specimens with a cycle threshold >35 tended to fail library preparation and were not sequenced, while samples with a cycle threshold between 32–35 tended to have poor genome coverage and we were unable to determine their SARS-CoV-2 classification.

**Supplementary Table 4. Distribution of samples that were unclassified for SARS-CoV-2 variant or failed library preparation**

|  | | | **Pre-October 2020** | | **October 2020 to April 2021** | | **May to November 2021** | |
| --- | --- | --- | --- | --- | --- | --- | --- | --- |
|  | **Failed library preparation** | **Unclassified** | **Failed library preparation** | **Unclassified** | **Failed library preparation** | **Unclassified** | **Failed library preparation** | **Unclassified** |
| **Any infection** | 8 | 30 | 0 | 1 | 7 | 26 | 1 | 3 |
| Placebo | 6 | 20 | 0 | 1 | 5 | 16 | 1 | 3 |
| Vaccine | 2 | 10 | 0 | 0 | 2 | 10 | 0 | 0 |
| **Mild/Moderate** | 5 | 18 | 0 | 1 | 4 | 15 | 1 | 2 |
| Placebo | 5 | 13 | 0 | 1 | 4 | 10 | 1 | 2 |
| Vaccine | 0 | 5 | 0 | 0 | 0 | 5 | 0 | 0 |
| **Moderate** | 2 | 4 | 0 | 0 | 1 | 4 | 1 | 0 |
| Placebo | 2 | 2 | 0 | 0 | 1 | 2 | 1 | 0 |
| Vaccine | 0 | 2 | 0 | 0 | 0 | 2 | 0 | 0 |

**Supplementary Table 5. Vaccine efficacy sensitivity analysis.** This table includes infections where the variant was unclassifiable. Infections pre-October 2020 were assigned a wild-type classification, between October 2020–April 2021 were assigned Beta classification and between May–November 2021 were assigned Delta classification.

| **Severity** | **SARS-CoV-2** | **Total cases** | **Placebo n = 960 Total FU time (days) = 244,140 Mean FU in days (SD) = 254 (71)** | | | **Vaccine n = 935 Total FU time (days) = 236,557 Mean FU in days (SD) = 253 (74)** | | | **VE (95% CI)** |
| --- | --- | --- | --- | --- | --- | --- | --- | --- | --- |
|  |  |  | **n (%)** | **Mean FU in months**  **(SD)** | **Incidence rate (person-days)** | **n (%)** | **Mean FU in months**  **(SD)** | **Incidence rate (person-days)** |  |
| **Any infection*** | WT | 13 | 12 (1.2) | 4.3 (3.1) | 17.9 | 1 (0.1) | 0.6 (2.2) | 1.5 | 91.4% (41.9, 99.8) |
|  | Beta | 132 | 73 (7.6) | 4.5 (1.5) | 109.1 | 59 (6.3) | 4.2 (NA) | 91.0 | 16.6% (-19.2, 41.8) |
|  | Delta | 26 | 22 (2.3) | 9.5 (1.2) | 32.9 | 4 (0.4) | 10 (0.7) | 6.2 | 81.2% (44.7, 95.3) |
| **Mild/ Moderate** | WT | 11 | 10 (1) | 4.2 (3.4) | 15 | 1 (0.1) | 0.6 (NA) | 1.5 | 89.7% (27.5, 99.8) |
|  | Beta | 89 | 51 (5.3) | 4.4 (1.6) | 76.2 | 38 (4.1) | 3.9 (1.8) | 58.6 | 23.1% (-19.4, 50.8) |
|  | Delta | 24 | 20 (2.1) | 9.6 (1.0) | 29.9 | 4 (0.4) | 10 (0.7) | 6.2 | 79.4% (38.4, 94.9) |

Abbreviations: CI, confidence interval; FU, follow-up; SD, standard deviation; VE, vaccine efficacy; WT, wild type; NA, not applicable.

^*^Includes symptomatic and asymptomatic infections.

**Supplementary Table 6. Summary of adverse events by system organ class and high-level** **preferred term**

| **N (%)** | **Overall** | **Placebo** | **Vaccine** |
| --- | --- | --- | --- |
| Any adverse event | 4640 | 2329 | 2311 |
| General disorders and administration site conditions | 1756 (82.6) | 836 (78.7) | 920 (86.5) |
| General system disorders NEC | 1634 (76.9) | 795 (74.9) | 839 (78.9) |
| Administration site reactions | 77 (3.6) | 23 (2.2) | 54 (5.1) |
| Body temperature conditions | 42 (2) | 16 (1.5) | 26 (2.4) |
| Therapeutic and nontherapeutic effects (excl toxicity) | 2 (<0.1) | 1 (<0.1) | 1 (<0.1) |
| Tissue disorders NEC | 1 (<0.1) | 1 (<0.1) | 0 (<0.1) |
| Eye disorders | 37 (1.7) | 21 (2) | 16 (1.5) |
| Ocular infections, irritations and inflammations | 33 (1.6) | 20 (1.9) | 13 (1.2) |
| Eye disorders NEC | 3 (0.1) | 1 (<0.1) | 2 (0.2) |
| Vision disorders | 1 (<0.1) | 0 (<0.1) | 1 (<0.1) |
| Metabolism and nutrition disorders | 33 (1.6) | 15 (1.4) | 18 (1.7) |
| Appetite and general nutritional disorders | 19 (0.9) | 8 (0.8) | 11 (1) |
| Electrolyte and fluid balance conditions | 8 (0.4) | 4 (0.4) | 4 (0.4) |
| Glucose metabolism disorders (incl diabetes mellitus) | 4 (0.2) | 1 (<0.1) | 3 (0.3) |
| Bone, calcium, magnesium and phosphorus metabolism disorders | 1 (<0.1) | 1 (<0.1) | 0 (<0.1) |
| Purine and pyrimidine metabolism disorders | 1 (<0.1) | 1 (<0.1) | 0 (<0.1) |
| Injury, poisoning and procedural complications | 30 (1.4) | 13 (1.2) | 17 (1.6) |
| Injuries NEC | 21 (1) | 9 (0.8) | 12 (1.1) |
| Injuries by physical agents | 4 (0.2) | 2 (0.2) | 2 (0.2) |
| Bone and joint injuries | 3 (0.1) | 1 (<0.1) | 2 (0.2) |
| Procedural related injuries and complications NEC | 2 (<0.1) | 1 (<0.1) | 1 (<0.1) |
| Reproductive system and breast disorders | 25 (1.2) | 11 (1) | 14 (1.3) |
| Menstrual cycle and uterine bleeding disorders | 16 (0.8) | 10 (0.9) | 6 (0.6) |
| Sexual function and fertility disorders | 4 (0.2) | 0 (<0.1) | 4 (0.4) |
| Reproductive tract disorders NEC | 2 (<0.1) | 1 (<0.1) | 1 (<0.1) |
| Vulvovaginal disorders (excl infections and inflammations) | 2 (<0.1) | 0 (<0.1) | 2 (0.2) |
| Uterine, pelvic and broad ligament disorders | 1 (<0.1) | 0 (<0.1) | 1 (<0.1) |
| Blood and lymphatic system disorders | 12 (0.6) | 6 (0.6) | 6 (0.6) |
| Anemias nonhemolytic and marrow depression | 5 (0.2) | 2 (0.2) | 3 (0.3) |
| Spleen, lymphatic and reticuloendothelial system disorders | 5 (0.2) | 4 (0.4) | 1 (<0.1) |
| Red blood cell disorders | 2 (<0.1) | 0 (<0.1) | 2 (0.2) |
| Ear and labyrinth disorders | 8 (0.4) | 2 (0.2) | 6 (0.6) |
| Aural disorders NEC | 7 (0.3) | 2 (0.2) | 5 (0.5) |
| External ear disorders (excl congenital) | 1 (<0.1) | 0 (<0.1) | 1 (<0.1) |
| Cardiac disorders | 6 (0.3) | 3 (0.3) | 3 (0.3) |
| Cardiac arrhythmias | 3 (0.1) | 1 (<0.1) | 2 (0.2) |
| Cardiac disorders, signs and symptoms NEC | 3 (0.1) | 2 (0.2) | 1 (<0.1) |
| Psychiatric disorders | 6 (0.3) | 5 (0.5) | 1 (<0.1) |
| Anxiety disorders and symptoms | 2 (<0.1) | 2 (0.2) | 0 (<0.1) |
| Depressed mood disorders and disturbances | 2 (<0.1) | 1 (<0.1) | 1 (<0.1) |
| Schizophrenia and other psychotic disorders | 1 (<0.1) | 1 (<0.1) | 0 (<0.1) |
| Sleep disorders and disturbances | 1 (<0.1) | 1 (<0.1) | 0 (<0.1) |
| Renal and urinary disorders | 5 (0.2) | 2 (0.2) | 3 (0.3) |
| Urinary tract signs and symptoms | 4 (0.2) | 1 (<0.1) | 3 (0.3) |
| Urolithiases | 1 (<0.1) | 1 (<0.1) | 0 (<0.1) |
| Immune system disorders | 4 (0.2) | 3 (0.3) | 1 (<0.1) |
| Allergic conditions | 4 (0.2) | 3 (0.3) | 1 (<0.1) |
| Respiratory, thoracic and mediastinal disorders | 681 (32) | 351 (33.1) | 330 (31) |
| Respiratory disorders NEC | 332 (15.6) | 173 (16.3) | 159 (15) |
| Respiratory tract signs and symptoms | 266 (12.5) | 132 (12.4) | 134 (12.6) |
| Upper respiratory tract disorders (excl infections) | 78 (3.7) | 44 (4.1) | 34 (3.2) |
| Pulmonary vascular disorders | 3 (0.1) | 2 (0.2) | 1 (<0.1) |
| Bronchial disorders (excl neoplasms) | 2 (<0.1) | 0 (<0.1) | 2 (0.2) |
| Endocrine disorders | 3 (0.1) | 3 (0.3) | 0 (<0.1) |
| Thyroid gland disorders | 3 (0.1) | 3 (0.3) | 0 (<0.1) |
| Hepatobiliary disorders | 3 (0.1) | 3 (0.3) | 0 (<0.1) |
| Hepatic and hepatobiliary disorders | 3 (0.1) | 3 (0.3) | 0 (<0.1) |
| Social circumstances | 3 (0.1) | 1 (<0.1) | 2 (0.2) |
| Legal issues | 3 (0.1) | 1 (<0.1) | 2 (0.2) |
| Pregnancy, puerperium and perinatal conditions | 2 (<0.1) | 1 (<0.1) | 1 (<0.1) |
| Abortions and stillbirth | 2 (<0.1) | 1 (<0.1) | 1 (<0.1) |
| Nervous system disorders | 654 (30.8) | 336 (31.6) | 318 (29.9) |
| Headaches | 489 (23) | 253 (23.8) | 236 (22.2) |
| Neurological disorders NEC | 101 (4.8) | 54 (5.1) | 47 (4.4) |
| Cranial nerve disorders (excl neoplasms) | 63 (3) | 28 (2.6) | 35 (3.3) |
| Peripheral neuropathies | 1 (<0.1) | 1 (<0.1) | 0 (<0.1) |
| Infections and infestations | 559 (26.3) | 276 (26) | 283 (26.6) |
| Infections ‒ pathogen unspecified | 468 (22) | 229 (21.6) | 239 (22.5) |
| Viral infectious disorders | 67 (3.2) | 36 (3.4) | 31 (2.9) |
| Fungal infectious disorders | 11 (0.5) | 5 (0.5) | 6 (0.6) |
| Bacterial infectious disorders | 10 (0.5) | 3 (0.3) | 7 (0.7) |
| Ectoparasitic disorders | 2 (<0.1) | 2 (0.2) | 0 (<0.1) |
| Helminthic disorders | 1 (<0.1) | 1 (<0.1) | 0 (<0.1) |
| Gastrointestinal disorders | 469 (22.1) | 243 (22.9) | 226 (21.3) |
| Gastrointestinal motility and defaecation conditions | 232 (10.9) | 117 (11) | 115 (10.8) |
| Gastrointestinal signs and symptoms | 217 (10.2) | 118 (11.1) | 99 (9.3) |
| Dental and gingival conditions | 7 (0.3) | 5 (0.5) | 2 (0.2) |
| Oral soft tissue conditions | 7 (0.3) | 2 (0.2) | 5 (0.5) |
| Gastrointestinal vascular conditions | 3 (0.1) | 1 (<0.1) | 2 (0.2) |
| Gastrointestinal hemorrhages NEC | 1 (<0.1) | 0 (<0.1) | 1 (<0.1) |
| Gastrointestinal inflammatory conditions | 1 (<0.1) | 0 (<0.1) | 1 (<0.1) |
| Gastrointestinal ulceration and perforation | 1 (<0.1) | 0 (<0.1) | 1 (<0.1) |
| Musculoskeletal and connective tissue disorders | 163 (7.7) | 102 (9.6) | 61 (5.7) |
| Muscle disorders | 63 (3) | 44 (4.1) | 19 (1.8) |
| Musculoskeletal and connective tissue disorders NEC | 53 (2.5) | 28 (2.6) | 25 (2.4) |
| Joint disorders | 45 (2.1) | 28 (2.6) | 17 (1.6) |
| Musculoskeletal and connective tissue deformities (incl intervertebral disc disorders) | 1 (<0.1) | 1 (<0.1) | 0 (<0.1) |
| Tendon, ligament and cartilage disorders | 1 (<0.1) | 1 (<0.1) | 0 (<0.1) |
| Skin and subcutaneous tissue disorders | 73 (3.4) | 54 (5.1) | 19 (1.8) |
| Epidermal and dermal conditions | 43 (2) | 33 (3.1) | 10 (0.9) |
| Skin appendage conditions | 27 (1.3) | 20 (1.9) | 7 (0.7) |
| Angioedema and urticaria | 3 (0.1) | 1 (<0.1) | 2 (0.2) |
| Investigations | 65 (3.1) | 22 (2.1) | 43 (4) |
| Cardiac and vascular investigations (excl enzyme tests) | 37 (1.7) | 11 (1) | 26 (2.4) |
| Physical examination and organ system status topics | 8 (0.4) | 1 (<0.1) | 7 (0.7) |
| Water, electrolyte and mineral investigations | 8 (0.4) | 5 (0.5) | 3 (0.3) |
| Hepatobiliary investigations | 5 (0.2) | 3 (0.3) | 2 (0.2) |
| Hematology investigations (incl blood groups) | 3 (0.1) | 1 (<0.1) | 2 (0.2) |
| Enzyme investigations NEC | 2 (<0.1) | 1 (<0.1) | 1 (<0.1) |
| Metabolic, nutritional and blood gas investigations | 1 (<0.1) | 0 (<0.1) | 1 (<0.1) |
| Microbiology and serology investigations | 1 (<0.1) | 0 (<0.1) | 1 (<0.1) |
| Vascular disorders | 43 (2) | 20 (1.9) | 23 (2.2) |
| Vascular hypertensive disorders | 40 (1.9) | 18 (1.7) | 22 (2.1) |
| Decreased and nonspecific blood pressure disorders and shock | 2 (<0.1) | 1 (<0.1) | 1 (<0.1) |
| Embolism and thrombosis | 1 (<0.1) | 1 (<0.1) | 0 (<0.1) |

Abbreviation: NEC, not elsewhere classified.

**Supplementary Table 7. Summary of serious adverse events by system organ class and high-level preferred term**

| **N (%)** | **Overall** | **Placebo** | **Vaccine** |
| --- | --- | --- | --- |
| Any serious adverse event | 58 | 24 | 34 |
| Injury, poisoning and procedural complications | 13 (0.6) | 4 (0.4) | 9 (0.8) |
| Injuries NEC | 7 (0.3) | 2 (0.2) | 5 (0.5) |
| Bone and joint injuries | 4 (0.2) | 1 (<0.1) | 3 (0.3) |
| Overdoses and underdoses NEC | 1 (<0.1) | 0 (<0.1) | 1 (<0.1) |
| Procedural related injuries and complications NEC | 1 (<0.1) | 1 (<0.1) | 0 (<0.1) |
| Hepatobiliary disorders | 2 (<0.1) | 2 (0.2) | 0 (<0.1) |
| Hepatic and hepatobiliary disorders | 2 (<0.1) | 2 (0.2) | 0 (<0.1) |
| Social circumstances | 2 (<0.1) | 1 (<0.1) | 1 (<0.1) |
| Legal issues | 2 (<0.1) | 1 (<0.1) | 1 (<0.1) |
| Immune system disorders | 1 (<0.1) | 0 (<0.1) | 1 (<0.1) |
| Allergic conditions | 1 (<0.1) | 0 (<0.1) | 1 (<0.1) |
| Investigations | 1 (<0.1) | 1 (<0.1) | 0 (<0.1) |
| Hepatobiliary investigations | 1 (<0.1) | 1 (<0.1) | 0 (<0.1) |
| Musculoskeletal and connective tissue disorders | 1 (<0.1) | 0 (<0.1) | 1 (<0.1) |
| Fractures | 1 (<0.1) | 0 (<0.1) | 1 (<0.1) |
| Nervous system disorders | 1 (<0.1) | 0 (<0.1) | 1 (<0.1) |
| Neurological disorders NEC | 1 (<0.1) | 0 (<0.1) | 1 (<0.1) |
| Reproductive system and breast disorders | 1 (<0.1) | 1 (<0.1) | 0 (<0.1) |
| Menstrual cycle and uterine bleeding disorders | 1 (<0.1) | 1 (<0.1) | 0 (<0.1) |
| Gastrointestinal disorders | 7 (0.3) | 3 (0.3) | 4 (0.4) |
| Exocrine pancreas conditions | 2 (<0.1) | 0 (<0.1) | 2 (0.2) |
| Gastrointestinal inflammatory conditions | 2 (<0.1) | 1 (<0.1) | 1 (<0.1) |
| Gastrointestinal conditions NEC | 1 (<0.1) | 1 (<0.1) | 0 (<0.1) |
| Gastrointestinal motility and defecation conditions | 1 (<0.1) | 1 (<0.1) | 0 (<0.1) |
| Gastrointestinal stenosis and obstruction | 1 (<0.1) | 0 (<0.1) | 1 (<0.1) |
| Infections and infestations | 7 (0.3) | 3 (0.3) | 4 (0.4) |
| Infections ‒ pathogen unspecified | 3 (0.1) | 2 (0.2) | 1 (<0.1) |
| Mycobacterial infectious disorders | 2 (<0.1) | 0 (<0.1) | 2 (0.2) |
| Bacterial infectious disorders | 1 (<0.1) | 0 (<0.1) | 1 (<0.1) |
| Viral infectious disorders | 1 (<0.1) | 1 (<0.1) | 0 (<0.1) |
| Pregnancy, puerperium and perinatal conditions | 6 (0.3) | 1 (<0.1) | 5 (0.5) |
| Abortions and stillbirth | 4 (0.2) | 1 (<0.1) | 3 (0.3) |
| Maternal complications of pregnancy | 2 (<0.1) | 0 (<0.1) | 2 (0.2) |
| General disorders and administration site conditions | 4 (0.2) | 0 (<0.1) | 4 (0.4) |
| Body temperature conditions | 2 (<0.1) | 0 (<0.1) | 2 (0.2) |
| General system disorders NEC | 2 (<0.1) | 0 (<0.1) | 2 (0.2) |
| Psychiatric disorders | 4 (0.2) | 2 (0.2) | 2 (0.2) |
| Schizophrenia and other psychotic disorders | 2 (<0.1) | 1 (<0.1) | 1 (<0.1) |
| Suicidal and self-injurious behaviors NEC | 2 (<0.1) | 1 (<0.1) | 1 (<0.1) |
| Respiratory, thoracic and mediastinal disorders | 4 (0.2) | 3 (0.3) | 1 (<0.1) |
| Pulmonary vascular disorders | 3 (0.1) | 2 (0.2) | 1 (<0.1) |
| Pleural disorders | 1 (<0.1) | 1 (<0.1) | 0 (<0.1) |
| Blood and lymphatic system disorders | 2 (<0.1) | 1 (<0.1) | 1 (<0.1) |
| Anemias nonhemolytic and marrow depression | 2 (<0.1) | 1 (<0.1) | 1 (<0.1) |
| Cardiac disorders | 2 (<0.1) | 2 (0.2) | 0 (<0.1) |
| Cardiac disorders, signs and symptoms NEC | 1 (<0.1) | 1 (<0.1) | 0 (<0.1) |
| Coronary artery disorders | 1 (<0.1) | 1 (<0.1) | 0 (<0.1) |

Abbreviation: NEC, not elsewhere classified.

**Supplementary Figure 1. Participant disposition**

2130 participants randomized*

1065 randomized to placebo

1065 randomized to AZD1222

989 received two doses of AZD1222

1034 received two doses of placebo

3 withdrew before first injection

27 missed second injection

1 mistakenly received AZD1222

2 withdrew before first injection

21 low-dose prime

23 low-dose booster

30 no booster

935 eligible AZD1222 participants

960 eligible placebo participants

72 COVID-19+ before 2 weeks post-booster

2 deaths before 2 weeks post-booster

54 COVID-19+ before 2 weeks
post-booster

*Updated from enrollment reported in [9], to exclude five participants who were randomized but withdrew before they received their first injection.

**Supplementary Figure 2.** Rolling mean number of total monthly SARS-CoV-2 infections through time. The color indicates the rolling mean of prevalence out of all samples for Alpha (light blue), Beta (green), Delta (dark blue) and unclassified (gray). The unclassified isolates were detected in individuals with viral loads too low (>32Ct) to get a definite strain classification.

Number of positive isolates

|  | 19A | 20A | 20B | 20D | Alpha | Beta | Delta | Unclassified |
| --- | --- | --- | --- | --- | --- | --- | --- | --- |
| 2020-06-01 |  |  | 6 | 1 |  |  |  |  |
| 2020-07-01 |  | 8 | 30 | 12 |  |  |  | 12 |
| 2020-08-01 |  |  | 6 |  |  |  |  | 12 |
| 2020-09-01 |  | 3 | 3 | 2 |  | 1 |  | 8 |
| 2020-10-01 |  | 2 | 2 | 1 |  | 5 |  | 2 |
| 2020-11-01 |  |  |  |  |  | 10 |  |  |
| 2020-12-01 |  |  | 1 |  |  | 34 |  |  |
| 2021-01-01 |  | 1 | 1 |  |  | 44 |  | 8 |
| 2021-02-01 |  | 1 | 3 |  |  | 12 |  | 17 |
| 2021-03-01 |  |  |  |  |  | 6 |  | 3 |
| 2021-04-01 |  |  |  |  |  | 2 |  |  |
| 2021-05-01 |  |  |  |  | 1 |  | 1 | 1 |
| 2021-06-01 | 1 | 1 |  | 3 | 2 | 4 | 65 | 6 |
| 2021-07-01 |  |  | 1 | 2 |  | 3 | 53 | 19 |
| 2021-08-01 |  |  |  | 1 |  |  | 13 | 7 |
| 2021-09-01 |  |  |  |  |  |  | 2 | 1 |

**Supplementary Figure 3.** Sensitivity analysis of cumulative incidence over time for any SARS-CoV-2 infection (includes both asymptomatic and symptomatic illness), due to wild-type (a), Beta (b) and Delta (c) variants. Infections pre-October 2020 were assigned a wild-type classification, between October 2020–April 2021 were assigned Beta classification and between May–November 2021 were assigned Delta classification.

a)

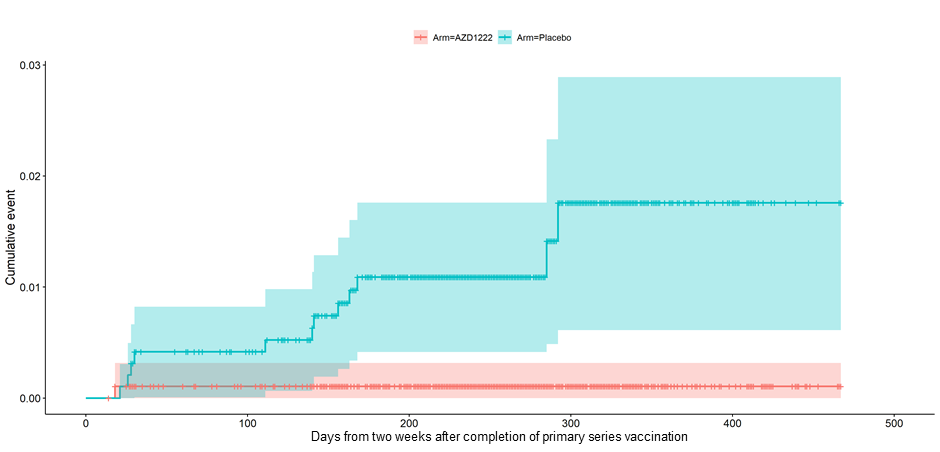

b)

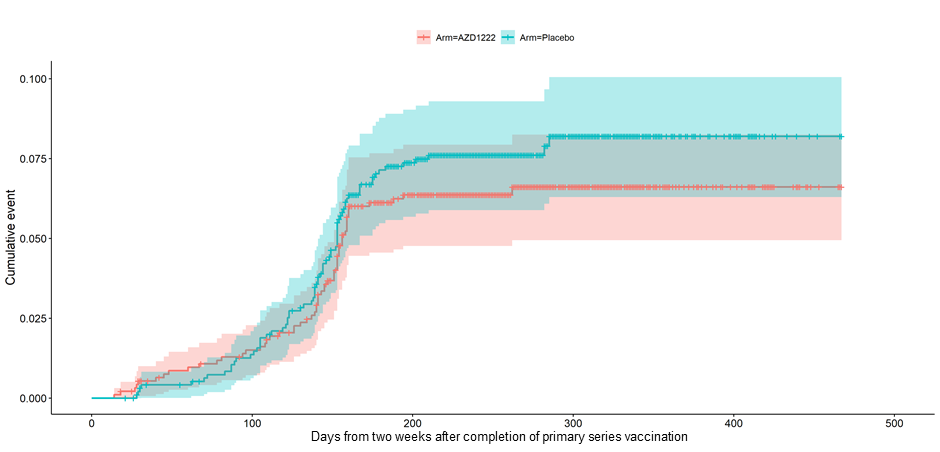

c)

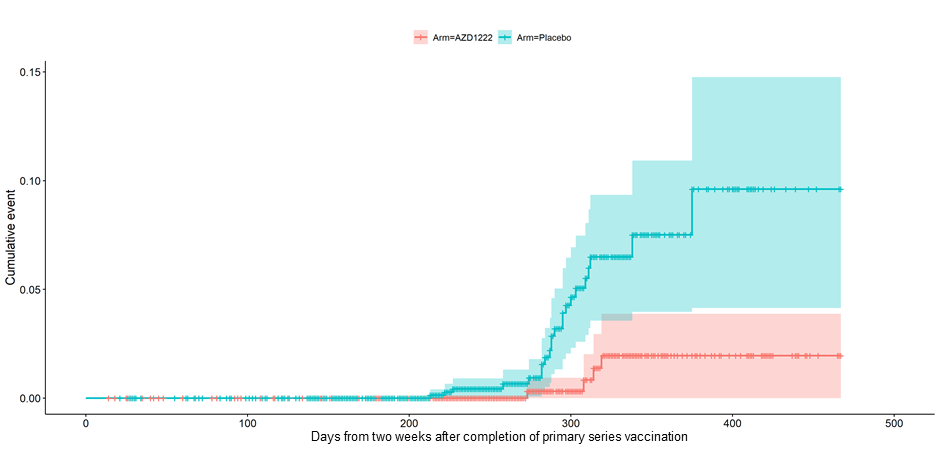
